# Profile of deaths mentioning ischemic and hemorrhagic stroke in Brazil: a population-based machine learning analysis

**DOI:** 10.1101/2025.03.05.25323460

**Authors:** Alessandro Rocha Milan de Souza, Letícia Martins Raposo, Glenda Corrêa Borges de Lacerda, Paulo Henrique Godoy

## Abstract

**Background and objective:** Brazil has the highest stroke rates in Latin America. The aim of this study was to investigate the profile of deaths mentioning stroke in Brazil between 2000 and 2019 and the relationship between multiple causes of death and stroke subtype.

**Methods:** Deaths mentioning stroke and other conditions were identified using individual death records from the country’s mortality information system (SIM). Strokes were grouped into the following subtypes according to the International Statistical Classification of Diseases and Related Health Problems 10th Revision (ICD-10) codes: ischemic stroke (IS); hemorrhagic stroke (HS); and unspecified stroke (US). Descriptive statistics were used to describe the distribution of stroke subtypes by demographic category, region and period. A decision tree model was built to identify the strongest factors distinguishing IS from HS.

**Results:** There were 2,459,742 deaths mentioning stroke. The following sociodemographic groups accounted for the highest proportion of overall deaths in their respective categories: the 60-79 age group (46.6%); white people (57%); married people (41.9%); and people with less than 3 years of education (62.1%). There was a progressive increase in the number of deaths mentioning stroke over the study period. The most common type of stroke was US, accounting for more than 60% of deaths. The most frequent group of causes of death in the subtype HS was hypertensive diseases (40.6%), while the most frequent group in subtypes IS and US was diseases of the respiratory system (48.30% and 42.30%, respectively). The decision tree analysis revealed that IS was more likely to occur in patients aged 60 years and over and in cases where respiratory diseases, endocrine diseases, arrhythmias, ischemic heart disease and heart failure were present. However, in younger patients without the previous mentioned diseases but where diseases of the nervous system were present HS was more frequent.

**Discussion:** The analysis of multiple causes of death enabled the identification of different diseases and factors associated with deaths mentioning IS, HS and US. The decision tree analysis identified the strongest factors distinguishing IS from HS. These findings can serve as guidance for defining the specific needs of each stroke subtype.

## Introduction

Stroke prevalence rates in Brazil vary between studies, regions and epidemiological contexts, ranging from 1.3% to 6.8%^1–4^.

Brazil has the highest stroke mortality rate in Latin America, with rates being higher among women. Although there has been a decline in stroke mortality in the country in recent decades, rates remain high^5^. A study by Souza et al. (in press) investigating trends in stroke mortality during the period 2000-2019 using data from the country’s mortality information system (SIM) reported an increase in cases of ischemic stroke (IS) from 2015, which contrasts with global trends^6^. However, the authors conclude that this increase may be related to improvements in reporting this cause of death in the SIM.

Strokes can be classified into two main groups: ischemic stroke (IS) and hemorrhagic stroke (HS). The latter is divided into intracerebral hemorrhage (ICH) and subarachnoid hemorrhage (SAH). IS is the most common type of stroke, accounting for around 65% of all cases worldwide, followed by ICH (26%) and SAH (9%)^7^. ICH mortality can be up to two times higher than IS mortality^8^. Risk factors differ according to stroke subtype, with systemic arterial hypertension and obesity being the leading factors for ICH and SAH and systemic arterial hypertension and diabetes for IS^9^.

Traditionally, the analysis of mortality has focused on the underlying cause of death. However, multiple causes of death, which include the underlying cause and contributing causes, provide a more complete picture of the process leading to death^10^.

The analysis of multiple causes helps capture the true magnitude of underlying conditions that may otherwise be hidden when only the underlying cause is considered, providing valuable information for the development of prevention strategies^11^. Considering the complexity of interactions between associated conditions and the sociodemographic factors that influence stroke mortality, this study investigated the profile of deaths mentioning stroke in Brazil between 2000 and 2019 and the relationship between multiple causes of death and stroke subtype.

## Materials and Methods

### Data Source

National mortality data for the period 2000-2019 were obtained from the SIM using R’s microdatasus package^12^.

### Identification of Causes of Death

Deaths mentioning stroke were identified using individual death records from the SIM, along with the other conditions and factors mentioned on the death certificate. Strokes were grouped into the following subtypes according to the International Statistical Classification of Diseases and Related Health Problems 10th Revision (ICD-10) codes: a) Ischemic stroke (IS): codes I63, I67.3, I67.8, I69.3; b) Hemorrhagic stroke (HS): codes I60, I61, I62, I69.0, I69.1, I69.2; and c) Stroke not specified as hemorrhagic or ischemic (US): codes I64, I67.9, I69.4, I69.8.

Cases where more than one stroke subtype was mentioned (approximately 2% of cases) were excluded to avoid ambiguity. Death records of individuals aged under 19 and where the year of death was missing were also excluded.

### Analysis of Multiple Causes

Multiple causes were defined as all causes mentioned on the death certificate, without distinguishing between underlying and contributing causes, as proposed by Santo^10^.

The ICD-10 codes for the other causes of death related to the stroke subtypes were grouped according to chapter. To facilitate data analysis and interpretation, the most frequent chapter (Chapter IX - Diseases of the circulatory system) was divided into the following groups: hypertensive diseases (I10 to I15); ischemic heart diseases (I20 to I25); heart failure (I50); cardiac arrhythmias (I48 to I49); peripheral vascular diseases (I70 to I99); and other diseases of the circulatory system not elsewhere classified (I00 to I09, I26 to I46, I51 to I69), excluding codes specifically related to the three stroke subtypes in code range I60 to I69.

Groups with a frequency of less than 1% were excluded. The chapter “Symptoms, signs and abnormal clinical and laboratory findings, not elsewhere classified ” (R00-R99) was also excluded because it does not include diseases and the codes in this group may be regarded as garbage codes.

### Final Dataset

The final dataset included the following variables: sex (female, male); race/skin color (yellow, white, indigenous, brown, black); marital status (single, consensual union, married, legally separated, widowed); education level (no education, 1-3 years, 4-7 years, 8-11 years, 12 years or more); place of death (home, hospital, other type of health facility, public thoroughfare, other); age group (19-39, 40-59, 60-79, 80 and over); region (North, Northeast, Midwest, Southeast, South); stroke subtype (IS, HS, US) and other causes of death mentioned on the death certificate.

### Statistical Analysis

Descriptive statistics were used to describe the distribution of stroke subtypes (IS, HS and US) by demographic category, region and period. The national and regional prevalence of each stroke subtype was also estimated.

### Decision Tree

To identify the strongest factors distinguishing IS from HS, we built a decision tree model from the sample of death records mentioning IS and HS. Decision trees provide a clear visual representation of decision-making processes and are an ideal method for healthcare professionals because they facilitate the interpretation and practical application of clinical data. They are particularly useful for capturing complex nonlinear relationships between variables, providing an effective tool for making informed medical decisions. The analysis was performed using the CART (Classification and Regression Tree) algorithm^13^, implemented using the rpart package in R^14^.

Decision trees sequentially apply a set of rules that split a predictor variable into a binary response. The splitting criterion is based on the concept of “purity”, where a node is pure if all its elements belong to a single class. When a node is impure, the algorithm splits the node to minimize impurity.

For model assessment and validation purposes, the dataset was divided into training and test sets using a 70/30 split. The complexity parameter (cp) was set at values ranging from 0.001 to 0.1 using the caret package train function^15^ in R and five-fold cross-validation and area under the ROC curve (AUC-ROC) as an evaluation metric. The ideal “cp ” value for the decision tree model was 0.001. This parameter is essential to prune decision tree complexity and avoid overfitting, controlling cost-complexity.

The graphical representation of the decision tree was built using the rpart.plot package^16^ in R. Model performance was assessed using the test set based on the AUC-ROC and interpreted following the guidelines proposed by Hosmer and Lemeshow^17^: ROC = 0.5 is considered no better than chance; 0.6-0.69 indicates poor discrimination; 0.7-0.79 indicates acceptable (reasonable) discrimination; 0.8-0.89 indicates good (excellent) discrimination; and 0.9-1.0 indicates outstanding discrimination.

## Results

There were 2,459,742 deaths mentioning stroke between 2000 and 2019. The period that accounted for the largest proportion of deaths mentioning stroke was 2015-2019 (26.5%). The regions with the highest and lowest number of deaths mentioning stroke were the Southeast (1,110,449; 45.1%) and North (126, 764; 5.2%), respectively. The following sociodemographic groups accounted for the highest proportion of overall deaths in their respective categories: the 60-79 age group (46.6%); white people (57%); married people (41.9%); and people with less than 3 years of education (62.1%). Men accounted for 50.7% of overall deaths. The most common place of death was a hospital (78%), followed by at home (17.1%) (Table 1). The most common type of missing data was education (28.61% of records), followed by marital status (7.4%), race (7.24%), place of death (0.13%) and sex (0.01%).

**Table 1.** Sociodemographic characteristics of deaths mentioning stroke (overall and by subtype)

|  | OVERALL | STROKE SUBTYPE |  |  |
| --- | --- | --- | --- | --- |
| Period and sociodemographic characteristic | N = 2,459,742<sup>1</sup> | Hemorrhagic<br>N = 472,052<sup>1</sup> | Ischemic<br>N = 470,401<sup>1</sup> | Unspecified<br>N = 1,517,289<sup>1</sup> |
| **Period** |  |  |  |  |
| 2000-2004 | 567,101 (23.1%) | 103,235 (21.9%) | 93,021 (19.8%) | 370,845 (24.4%) |
| 2005-2009 | 605,534 (24.6%) | 111,169 (23.6%) | 103,603 (22.0%) | 390,762 (25.8%) |
| 2010-2014 | 635,649 (25.8%) | 122,400 (25.9%) | 121,474 (25.8%) | 391,775 (25.8%) |
| 2015-2019 | 651,458 (26.5%) | 135,248 (28.7%) | 152,303 (32.4%) | 363,907 (24.0%) |
| **Region** |  |  |  |  |
| Midwest | 145,622 (5.9%) | 32,010 (6.8%) | 31,079 (6.6%) | 82,533 (5.4%) |
| Northeast | 643,693 (26.2%) | 107,402 (22.8%) | 88,496 (18.8%) | 447,795 (29.5%) |
| North | 126,764 (5.2%) | 24,912 (5.3%) | 16,748 (3.6%) | 85,104 (5.6%) |
| Southeast | 1,110,449(45.1%) | 236,795 (50.2%) | 234,149 (49.8%) | 639,505 (42.1%) |
| South | 433,214 (17.6%) | 70,933 (15.0%) | 99,929 (21.2%) | 262,352 (17.3%) |
| **Age group (years)** |  |  |  |  |
| 19-39 | 76,438 (3.1%) | 46,227 (9.8%) | 9,306 (2.0%) | 20,905 (1.4%) |
| 40-59 | 409,258 (16.6%) | 161,417 (34.2%) | 62,395 (13.3%) | 185,446 (12.2%) |
| 60-79 | 1,145,977 (46.6%) | 187,589 (39.7%) | 226,729 (48.2%) | 731,659 (48.2%) |
| ≥ 80 | 828,069 (33.7%) | 76,819 (16.3%) | 171,971 (36.6%) | 579,279 (38.2%) |
| **Sex** |  |  |  |  |
| Female | 1,212,314 (49.3%) | 233,129 (49.4%) | 238,844 (50.8%) | 740,341 (48.8%) |
| Male | 1,247,168 (50.7%) | 238,877 (50.6%) | 231,517 (49.2%) | 776,774 (51.2%) |
| DC<sup>2</sup> | 260 (0.01%) | 46 | 40 | 174 |
| **Race/skin color** |  |  |  |  |
| Yellow | 16,147 (0.7%) | 3,742 (0.9%) | 3,261 (0.7%) | 9,144 (0.7%) |
| White | 1,301,687 (57.0%) | 241,956 (55.3%) | 280,430 (63.5%) | 779,301 (55.6%) |
| Indigenous | 3,742 (0.2%) | 690 (0.2%) | 531 (0.1%) | 2,521 (0.2%) |
| Brown | 749,505 (32.8%) | 151,030 (34.5%) | 120,789 (27.3%) | 477,686 (34.1%) |
| Black | 210,581 (9.2%) | 39,899 (9.1%) | 36,803 (8.3%) | 133,879 (9.5%) |
| DC<sup>2</sup> | 178,080 (7.24%) | 34,735 | 28,587 | 114,758 |
| **Marital Status** |  |  |  |  |
| Single | 453,161 (19.9%) | 111,850 (25.7%) | 76,770 (17.5%) | 264,541 (18.8%) |
| C Union<sup>3</sup> | 34,925 (1.5%) | 8,808 (2.0%) | 5,607 (1.3%) | 20,510 (1.5%) |
| Married | 953,753 (41.9%) | 196,439 (45.2%) | 180,205 (41.0%) | 577,109 (41.1%) |
| Separated | 113,147 (5.0%) | 29,609 (6.8%) | 23,975 (5.5%) | 59,563 (4.2%) |
| Widowed | 723,502 (31.8%) | 88,254 (20.3%) | 153,305 (34.9%) | 481,943 (34.3%) |
| DC<sup>2</sup> | 181,254(7.4%) | 37,092 | 30,539 | 113,623 |
| **Education level** |  |  |  |  |
| None | 503,908 (28.7%) | 52,763 (16.2%) | 80,089 (23.8%) | 371,056 (33.9%) |
| 1-3 y | 587,334 (33.4%) | 98,464 (30.2%) | 117,877 (35.1%) | 370,993 (33.9%) |
| 4-7 y | 394,355 (22.5%) | 89,701 (27.5%) | 80,705 (24.0%) | 223,949 (20.5%) |
| 8-11 y | 188,750 (10.7%) | 58,261 (17.9%) | 39,862 (11.9%) | 90,627 (8.3%) |
| ≥12 y | 81,621 (4.6%) | 26,748 (8.2%) | 17,339 (5.2%) | 37,534 (3.4%) |
| DC<sup>2</sup> | 703,774(28.61%) | 146,115 | 134,529 | 423,130 |
| **Place of death** |  |  |  |  |
| At home | 419,374 (17.1%) | 26,532 (5.6%) | 53,001 (11.3%) | 339,841 (22.4%) |
| Hospital | 1,917,378 (78.0%) | 422,563 (89.6%) | 399,274 (84.9%) | 1,095,541 (72.3%) |
| Other<sup>4</sup> | 73,706 (3.0%) | 9,861 (2.1%) | 11,493 (2.4%) | 52,352 (3.5%) |
| Pub<br>thoroughfare<sup>5</sup> | 14,438 (0.6%) | 7,191 (1.5%) | 901 (0.2%) | 6,346 (0.4%) |
| Other | 31,781 (1.3%) | 5,427 (1.2%) | 5,400 (1.1%) | 20,954 (1.4%) |
| DC<sup>3</sup> | 3,065(0.13%) | 478 | 332 | 2,255 |

<sup>1</sup>Total number
<sup>2</sup>Unknown
<sup>3</sup>Consensual union
<sup>4</sup>Other type of health facility
<sup>5</sup>Public thoroughfare

The most common type of stroke was US, accounting for 62% (1,517,289) of all deaths mentioning stroke, followed by HS (19.1%; 472,052) and IS (18.9%; 470,401). The number of deaths mentioning IS and HS increased over the study period, while the number mentioning US remained relatively stable. HS accounted for a higher number of deaths than IS in all regions except the South, with the latter region accounting for 21.2% of all deaths mentioning IS. In the Northeast and North, US accounted for a larger proportion of deaths (29.5% and 5.6%, respectively) than IS and HS. Younger individuals (aged up to 59 years) accounted for 44% of all deaths mentioning HS (compared to 56% in the older age groups) and only 15.3% and 13.6%, respectively, of deaths mentioning IS and US (compared to 84.8% and 86.4%, respectively, in the older age groups). Women accounted for a higher proportion of deaths mentioning IS than men (50.8% versus 49.2%).

White people accounted for the highest proportion deaths overall and by subtype, followed by brown people, with whites representing 63.5% of all deaths mentioning IS and brown people accounting for 34.5% of all deaths mentioning HS. Indigenous people accounted for the smallest proportion of over all deaths (0.2%). In the category marital status, married people accounted for the largest proportion of deaths across all stroke subtypes. The proportion of deaths represented by widows was highest in the IS and US subtypes (34.9% and 34.3%), while the share of deaths represented by single people was highest in the HS subtype (25.7%). With regard to education level, the results show that the group with the highest level of education accounted for the lowest proportion of deaths overall and across all stroke subtypes. In the subtype IS, people with no education accounted for 23.8% of deaths, while those with more than 12 years of education represented only 5.2%. The most common place of death was a hospital, followed by at home, which accounted for 22.4% of deaths mentioning US (Table 1).

The most frequently mentioned group of causes of death among overall deaths mentioning stroke was diseases of the respiratory system (40.2%), followed by hypertensive diseases (38%), infectious diseases (18.1%) and endocrine diseases (17.2%). The most frequent group among deaths mentioning HS was hypertensive diseases (40.6% of cases), while the most frequent group among deaths mentioning IS and US was diseases of the respiratory system (48.30% and 42.30%, respectively). The frequency of endocrine diseases, infectious diseases, arrhythmia, ischemic heart disease and heart failure was 19.2%, 24.1%, 7.6%, 6.7% and 6.2%, respectively, among deaths mentioning IS, and 9.2%, 12.1%, 2.4%, 2.5% and 1.6%, respectively, among deaths mentioning HS. In contrast, the frequency of hypertensive diseases, diseases of the nervous system and cerebrovascular diseases not associated with brain injury was 40.6%, 17.2% and 3.9% among deaths mentioning HS and 34.6%, 7.9% and 2.2%, among deaths mentioning IS. The frequencies of the groups of causes of death related to US and IS were similar, except for arrhythmias, peripheral vascular diseases and hypertensive diseases, where similarities were found between US and HS (Figure 1).

**Figure 1.**
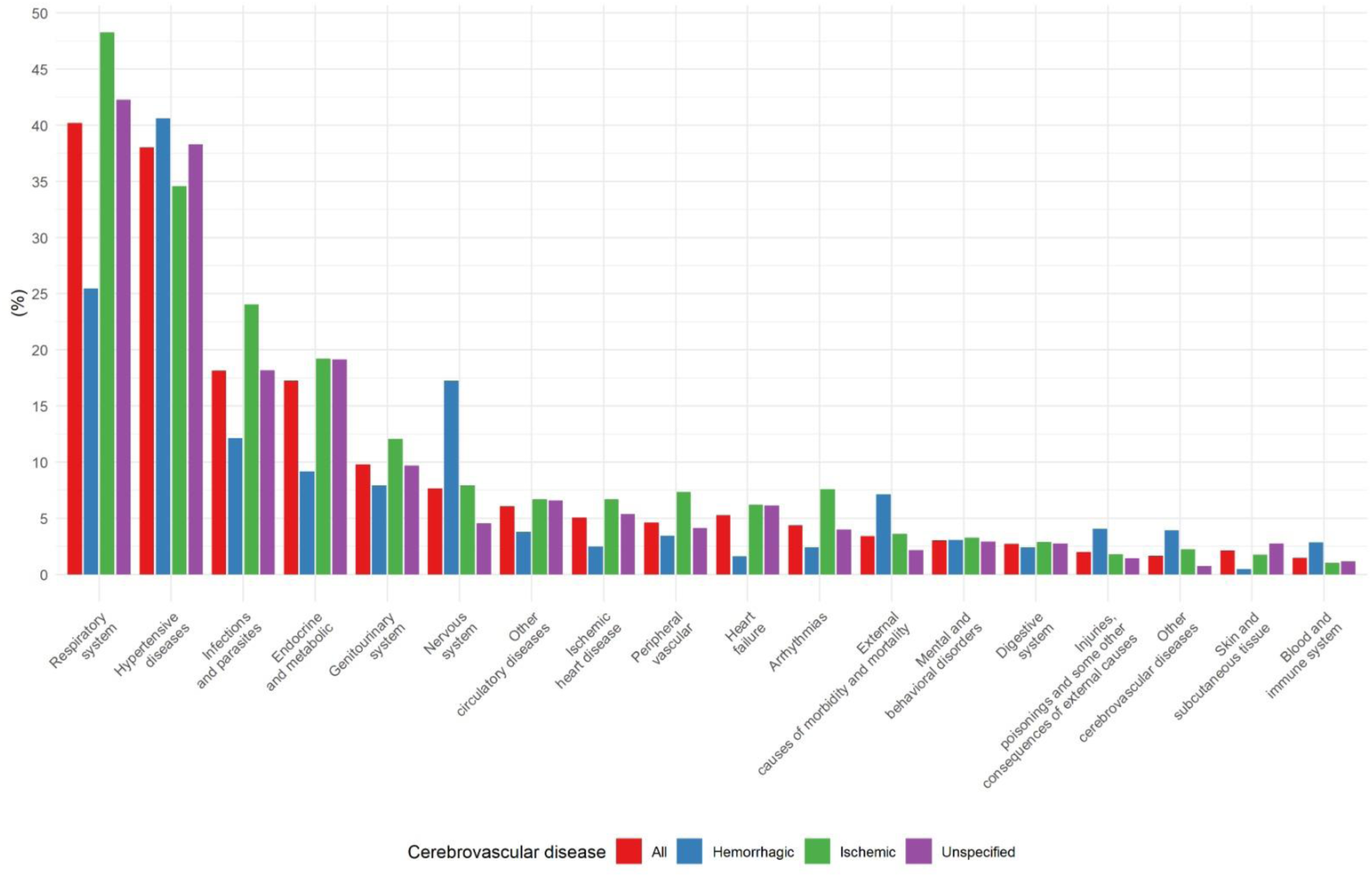
Frequency of groups of causes of death in deaths mentioning stroke (overall and by subtype).

Figure 2 shows the frequency of groups of causes of death among deaths mentioning stroke by subtype and region. The most frequent group among deaths mentioning HS and IS was hypertensive diseases, followed by diseases of the respiratory system, across all regions. Hypertensive diseases was the most frequent group in the North across all stroke subtypes. Among the subtypes HS and IS, the frequency of diseases of the respiratory system and nervous system was highest in the Midwest (27.5% and 20.9% and 52.8% and 9.1%, respectively). In the subtype HS, the frequency of external causes of morbidity and mortality (15.10%) and injury, poisoning and certain other consequences of external causes (11.2%) was highest in the North (Figure 2).

**Figure 2.**
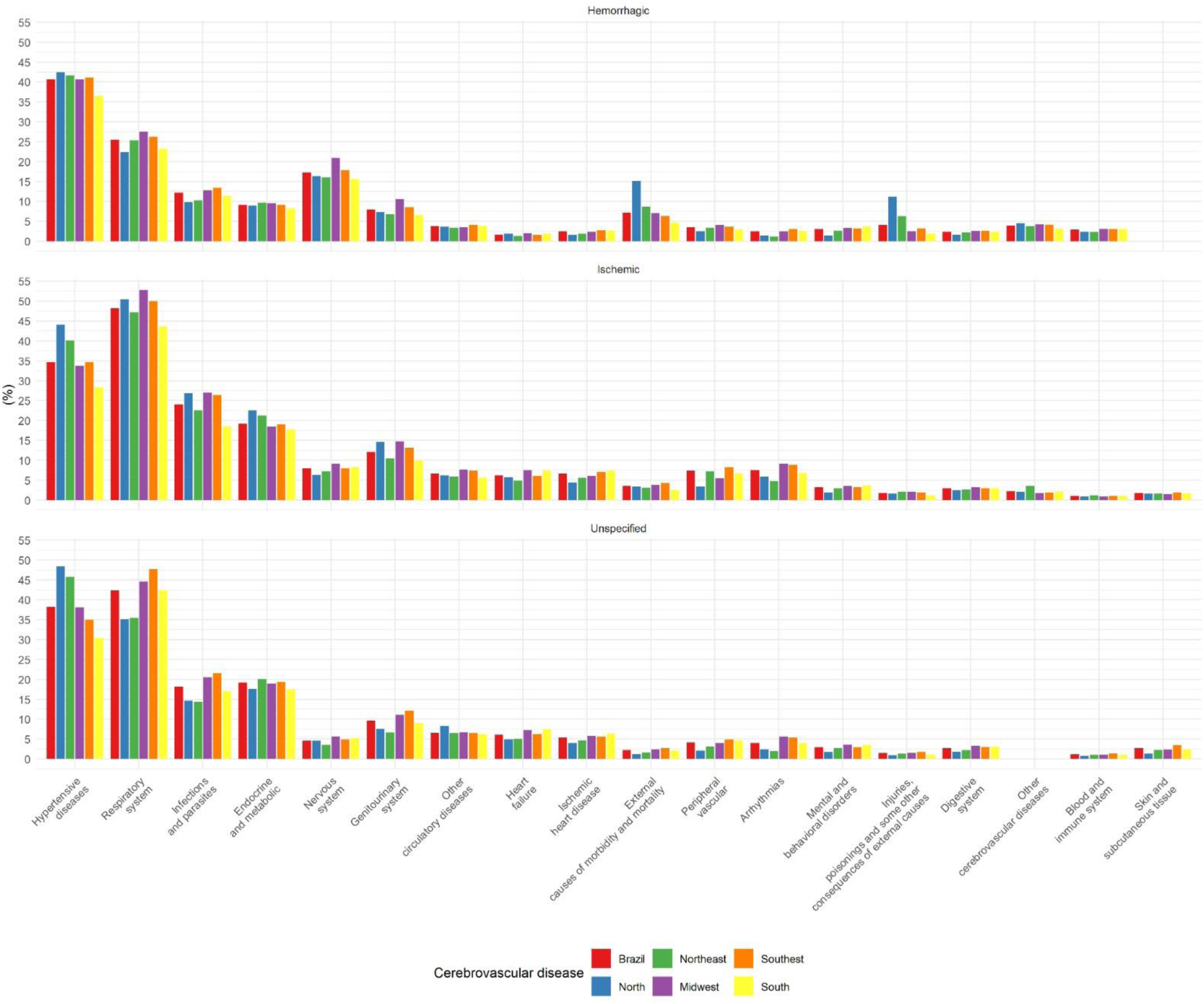
Frequency of groups of causes of death among deaths mentioning stroke by subtype and region.

## DECISION TREE ANALYSIS

The decision tree model (Figure 3) classifying deaths mentioning HS and IS was built from a dataset containing 442,451 observations.

**Figure 3.**
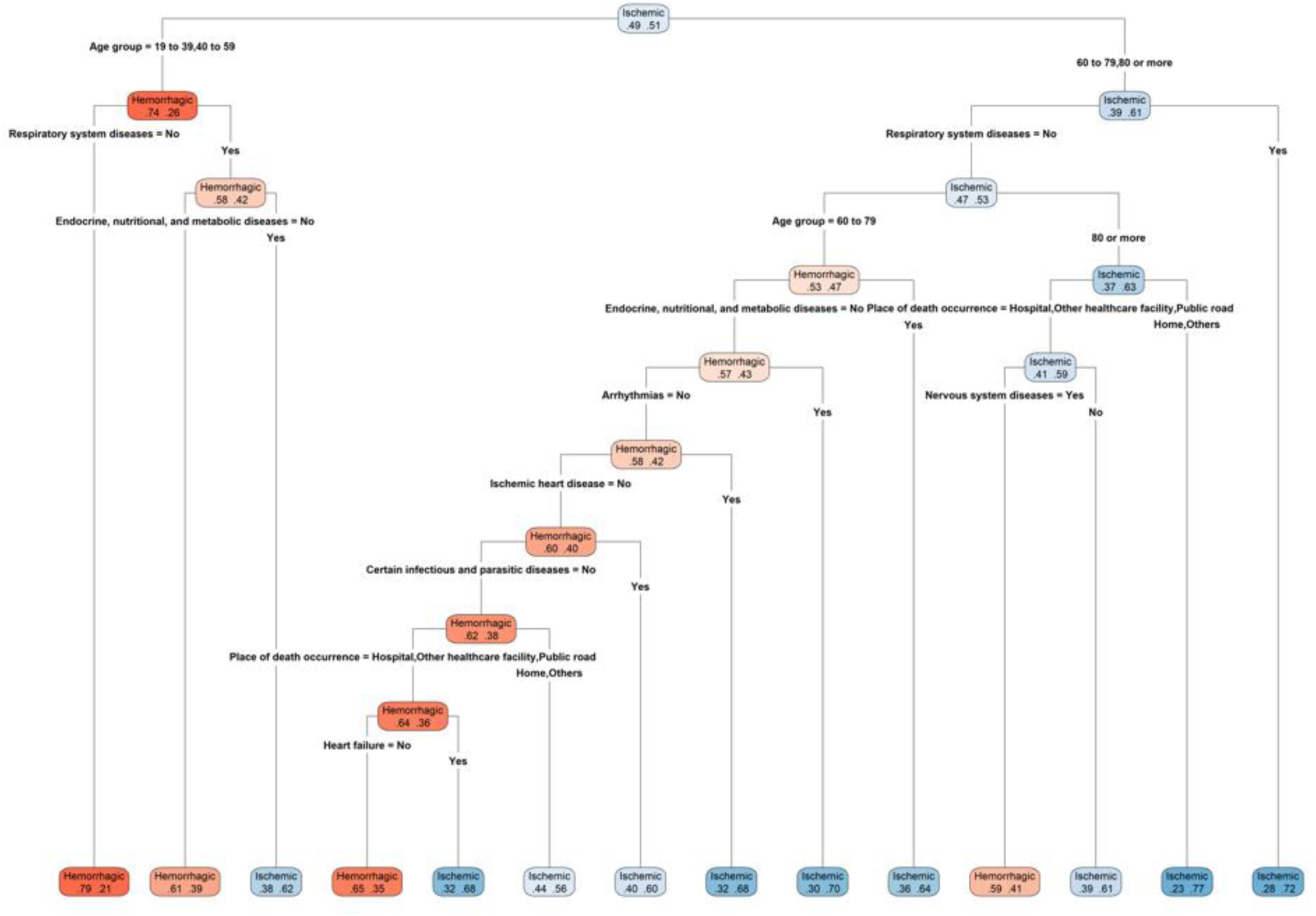
Decision tree for hemorrhagic and ischemic stroke. The percentages on the left represent hemorrhagic stroke and the colors represent magnitude.

The model’s AUC value was 0.727 (95% CI 0.724 – 0.729), indicating reasonable performance. At the root node, the deaths were distributed as follows: HS = 49% and IS = 51%. The first split was based on age, with 74% of individuals aged between 19 and 59 years being classified as HS. Within this subgroup, the proportion of deaths mentioning HS was considerably higher in cases not mentioning diseases of the respiratory system than in cases where this group was present (79% versus 58%). When cases were split based on the presence of endocrine, nutritional and metabolic diseases, the proportion of deaths mentioning HS is considerably higher in cases not mentioning these diseases than in cases where this group was present (61% versus 38%) (Figure 3). In the age group 60 years and over, most cases were classified as IS (61%). This proportion was even higher among deaths mentioning respiratory diseases (72%). When cases without the presence of respiratory diseases were split by age group (60-79 and 80 and over), most of the deaths in the older group were classified as IS (63%). This node was then split into place of death, with most of the cases of death in a hospital, other health facility or public thoroughfare without the presence of diseases of the nervous system being classified as IS (61%). In contrast, most of the cases with the presence of this group were classified as HS (59%). Most of the cases where place of death was at home or other were classified as IS (77%) (Figure 3).

In the 60-79 age group without the presence of respiratory diseases 53% of the deaths were classified as HS. Within this subgroup, 64% of the cases with the presence of endocrine, nutritional and metabolic diseases mentioned IS. In cases not mentioning these diseases but mentioning arrhythmias, this proportion increased to 70%. In cases not mentioning arrhythmia but with the presence of ischemic heart disease, 68% of the deaths were classified as IS. In contrast, in cases not mentioning ischaemic heart disease but with the presence of certain infectious and parasitic diseases the frequency of IS fell to 60%. This rate was even lower (56%) in cases where the place of death was at home or other without the presence of certain infectious and parasitic diseases. In deaths occurring in a hospital, other type of health facility or public thoroughfare with the presence of heart failure the frequency of IS increased to 68%. In contrast, in these cases without the presence of heart failure the predominant stroke subtype was HS (65%) (Figure 3).

## Discussion

The period with the largest proportion of deaths mentioning stroke was 2015-2019. A previous study investigating trends in stroke mortality based on underlying cause (in press) reported an upward trend in deaths from stroke over this period^6^. The authors attributed this increase to a decrease in US over the period explained by improvements in SIM data quality.

The Southeast continues to account for the highest number of deaths in Brazil. Higher mortality rates in more developed states may be explained by the fact that chronic conditions have a greater influence on the mortality profile in these more populous regions^18^.

US remains the most common type of stroke despite a decline in the number of ill-defined causes of death across the country^19^. In the Northeast and North, US accounted for a higher proportion of deaths mentioning stroke (29.5% and 5.6%, respectively) than IS and HS. Garritano et al. also found that US accounted for a higher proportion of deaths than IS and HS, representing more than 15% of deaths in older adults in the North^20^, while Jorge et al. reported that US accounted for over 20% of stroke deaths in the North and Northeast^21^. Key problems in these regions include poor access to health care related to their huge geographic area and cultural factors influencing health behavior^22^.

Our findings show that younger adults account for a larger proportion of deaths mentioning HS, while older adults account for a larger proportion of cases mentioning IS and US. These findings corroborate the results of a study by De Moraes et al., who found that 76% of deaths from HS in the South and Southeast of Brazil were among younger patients (10-49 years)^23^.

Our results show that the number of white people afflicted by stroke was more than that of black and brown people combined across all stroke subtypes. This can be explained by the fact that the population in the regions that account for the highest numbers of deaths from stroke (the Southeast and South) is predominantly white^24^. However, the international literature reports that stroke incidence^25^, the prevalence of risk factors such hypertension and diabetes, and levels of CVD biomarkers such as lipoprotein A^26^ tend to be higher among black people. In addition, this group also experiences inequalities across various social determinants of health, tending to have lower socioeconomic status and education level and poorer access to health care than white people^27^. In a study of race-adjusted stroke deaths in Brazil in 2010 based on underlying cause, Lotufo and Bensenor^28^found that mortality was higher among black people. The difference between our results and the findings of the above study may be explained by the fact that the current study investigated stroke deaths based on multiple causes rather than just underlying causes. Indigenous people accounted for the lowest proportion of deaths in our sample, which is consistent with the findings of previous study^29^.

A previous study found that married people accounted for the highest proportion of deaths from stroke in the category marital status^30^.

The fact that the proportion of deaths is lower in individuals with a higher level of education may be explained by higher income and better access to goods and services among this group, including education and health care^31^.

The fact that a higher proportion of deaths mentioning US occurred at home may be explained by diagnostic challenges and the absence of a death verification service^32^. Challenges in defining death in non-healthcare settings were highlighted by Santo in a study investigating ill-defined deaths in Brazil in 2003, showing that 53.3% these deaths were unattended^33^. However, the fact that the proportion of deaths mentioning US in our study were in-hospital deaths may be partially explained by poor diagnosis skills, given that there was no significant change this percentage with the presence of a CT scan^34^.

The most frequent group of causes of death in deaths mentioning stroke at national level and across regions was diseases of the respiratory system, followed by hypertensive diseases, ill-defined causes, infectious diseases and endocrine diseases. A study by Santo and Pinheiro^35^ in São Paulo showed that the primary causes related to stroke deaths were respiratory diseases (41.8%), hypertension (37.8%) and ill-defined causes (26.3%). A study in the state of Paraná by Furukawa et al.^36^ reported that the leading causes of death related to stroke deaths were diseases of the circulatory system (52.2%), diseases of the respiratory system (31.1%) and ill-defined causes (27%), while an investigation in Belo Horizonte by Ishitani et al^37^ found that the main causes were hypertensive diseases (33.8%) and respiratory system diseases (28.1%).

Strokes have a high rate of complications, with prevalence of post-stroke dysphagia standing at around 40%^38^. Stroke patients with dysphagia have an 8.5-fold higher risk of death, a higher risk of pneumonia and higher in-hospital costs^39^. In contrast with the present study, which found that respiratory system diseases were more frequent in deaths mentioning IS, a meta-analysis by Banda et al. investigating the prevalence of dysphagia and risk of pneumonia and mortality in acute stroke patients found that prevalence of dysphagia was higher in patients with HS (OR 1.52 (95%CI, 1.13–2.07). Our findings show that pneumonia and respiratory failure account for 90% of the mentions in the group diseases of the respiratory system (data not shown).

Laurenti^40^ and Santo^15^ reported that primary arterial hypertension was related to stroke in 75.9% and 57.4% of deaths, respectively. In the present study, primary arterial hypertension was more frequent in deaths mentioning HS than in those mentioning IS (40.6% versus 34.6%). The INTERSTROKE study^41^ also found that the association between hypertension and stroke was stronger for HS than for IS.

While certain diseases were mentioned together with stroke on the death certificate, causal relationships cannot be inferred. Some of these diseases may be risk factors, while others may be complications related to the underlying cause^42^. Our findings suggest that this may be the case with primary hypertension and diseases of the respiratory system, which were the two most prevalent causes at national level and across regions.

Endocrine diseases, arrhythmias, ischemic heart disease and heart failure were more frequent in deaths mentioning IS than among those mentioning HS. In a study examining the relationship between vascular risk factors and stroke type in native black Africans, Owolabi and Agunloye^43^ found that the association between arrhythmias, heart failure and diabetes and stroke was stronger for IS than for HS.

Frequency of hypertensive diseases was highest in the North, followed by the Northeast, across all stroke subtypes. Although the 2019 National Health Program (PNS)^44^ shows that the North and Northeast are not the regions with the highest proportion of people diagnosed with hypertension, access to medications dispensed by the popular pharmacy was lowest in these regions (34.8% and 38% respectively, compared to 45% nationally). In addition, the percentage of hypertensive patients undergoing drug therapy for the condition is lower in these regions^45^. Studies of the prevalence of chronic diseases such as primary arterial hypertension show that rates are higher among economically disadvantaged groups^46^. Furthermore, in an ecological study investigating the relationship between socioeconomic indicators and cardiovascular disease mortality in 98 municipalities in Brazil, Ishitani et al.^40^ found an inverse correlation between death from cardiovascular diseases, cerebrovascular diseases and hypertensive diseases and income, and a direct association with poverty and precarious living conditions.

In the North, external causes of morbidity and mortality and injury, poisoning and certain other consequences of external causes were more frequent in deaths mentioning HS than in deaths mentioning IS. According to the 2019 National Health Survey (NHS), the proportion of individuals who drove a motor vehicle after drinking was 17.0% nationally, ranging from 14.8% in the South and Southeast to 23.4% in the North. This rate was higher in rural areas (22.5% compared to 16.2% in urban areas)^44^. The higher frequency of the group injury, poisoning and certain other consequences of external causes in the North may therefore be explained by higher rates of head trauma due to accidents in this region.

The decision tree revealed complex patterns in the classification of deaths mentioning HS and IS, revealing that the distinction between the two depends on complex interactions between age and the presence of respiratory diseases and other comorbidities. Age group was the strongest initial factor, with younger individuals (19-59 years) being classified predominantly as HS and older individuals (60 years and older) predominantly as IS. These findings are consistent with those of Bernal et al., who investigated the incidence of hospitalization and mortality due to stroke in young adults in the Southeast and South of Brazil, finding that HS accounted for 76% of the 78,123 hospitalisations due to stroke in this group between 2008-2018^23^.

In older adults, the frequency of IS was higher in cases with the presence of respiratory system diseases, while in younger adults the frequency of HS was higher in cases not mentioning this group of causes of death. In a study with 570 consecutive patients with IS treated in a tertiary stroke center in Switzerland by Arnold et al., dysphagia was diagnosed in 20.7% of the patients, increasing the chance of aspiration pneumonia. At 3-month follow-up, 13.3% of the patients with dysphagia died due to respiratory complications^39^. These findings may partially explain the higher frequency of IS in cases mentioning respiratory system diseases among older adults in our study.

Other strong factors included endocrine, nutritional and metabolic diseases, arrhythmias, and place of death. Cases mentioning endocrine, nutritional and metabolic diseases were predominantly IS, regardless of age group. These findings are consistent with the findings of a collaborative meta-analysis of 102 prospective studies showing that adjusted hazard ratios with diabetes was 2.27 (1.95-2.65) for IS and 1.56 (1.19-2.05) for HS^47^. In patients aged over 60, arrhythmias, ischaemic heart disease and heart failure were more frequent in deaths mentioning IS. The Framingham study^48^ showed that there was a fivefold excess of stroke when atrial fibrillation was present, while Adelborg et al. found an increase in the stroke rate ratio in patients with heart failure^49^. Using autopsy data, Gongora-Rivera et al.^50^ found that myocardial infarction was present in 40.8% of 251 patients who died from IS. These findings may explain the higher frequency of arrhythmias, ischaemic heart disease and heart failure in deaths mentioning IS.

Place of death was also a strong factor, with deaths at home being predominantly IS, especially among older adults. It is therefore possible that most US deaths at home among individuals aged 80 and over are actually IS.

The technique used by the current study to analyze the association between multiple causes of death (machine learning decision tree) is different to that employed by Santo & Pinheiro^35^. The machine learning decision tree showed how the interaction of multiple factors can influence the likelihood of a stroke being complex (ischemic) or hemorrhagic.

This study provides some important insights into the profile of deaths from stroke in Brazil. Model performance suggests acceptable discriminant validity, suggesting that the decision tree was effective in identifying relevant patterns and important associations, despite the limitations of the data. While secondary data from the SIM have some limitations, including recording errors and underreporting, the system provides comprehensive representative data encompassing millions of records over a period of two decades. In addition, while our data is limited to sociodemographic variables and mentions of other conditions, we explored interactions between factors, providing important information that can serve as guidance for the design of public health interventions. Our findings make an important contribution to existing knowledge on stroke mortality, underlining the importance of prevention policies that focus on key risk factors and diseases for stroke death and reinforcing the need for further research investigating other clinical variables to enhance risk prediction and preventive interventions.

## Conclusions

The analysis of multiple causes enabled the identification of all causes related to deaths mentioning ischemic and hemorrhagic stroke, providing valuable insights into the profile of stoke deaths, highlighting important contributing conditions and offering important inputs to help shape care planning. The decision tree revealed factors, conditions and diseases with a stronger association with IS and HS, which can serve as guidance for the redistribution of unspecified strokes.

## Data Availability

I, the principal author, have full access to the data used in the analyses in the manuscript and I take full responsibility for the data, the analyses and interpretation, and the conduct of the research; I have the right to publish any and all data, separate and apart from the guidance of any sponsor.

